# Transcriptomic profile of normal breast tissue post-mifepristone treatment: secondary outcomes of a randomized controlled trial

**DOI:** 10.1101/2024.03.08.24303979

**Authors:** Deborah Utjés, Nageswara Rao Boggavarapu, Mohammed Rasul, Isabelle Koberg, Alexander Zulliger, Sakthivignesh Ponandai-Srinivasan, Carolina von Grothusen, Parameswaran Grace Lalitkumar, Kiriaki Papaikonomou, Twana Alkasalias, Kristina Gemzell-Danielsson

## Abstract

Progesterone receptor antagonism is gaining attention due to progesterone’s recognized role as a major mitogen in breast tissue. Limited but promising data suggest the potential efficacy of antiprogestins in breast cancer prevention. The present study presents secondary outcomes from a randomized controlled trial and examine changes in breast mRNA expression following mifepristone treatment in healthy women. We analyzed 32 paired breast biopsies from 16 healthy premenopausal women at baseline and after two months of mifepristone treatment. In total, twenty-seven differentially expressed genes were identified, with enriched biological functions related to extracellular matrix remodeling. Notably, the altered gene signature induced by mifepristone *in vivo* was rather similar to the *in vitro* signature. Furthermore, this expression gene signature was associated with breast carcinogenesis and significantly correlated with progesterone receptor expression status in breast cancer, as validated in The Cancer Genome Atlas dataset using the R2 platform. The present study is the first to explore the breast transcriptome following mifepristone treatment in healthy breast tissue *in vivo*, enhancing the understanding of progesterone receptor antagonism and its potential protective effect against breast cancer by investigating its action in healthy breast tissue.

## Introduction

The majority of breast cancers are related to reproductive factors (1), implicating endogenous cyclic hormonal exposure affecting breast tumorigenesis. Progesterone has emerged as a major mitogen since proliferation of breast epithelial cells occur during the progesterone-dominated luteal phase (2–4). Among different epithelial cell subtypes, the luminal progenitors serve as breast cancer precursor cell (5). Progesterone acts primarily via a paracrine mechanism to stimulate proliferation of the dominating progesterone receptor (PR)-negative breast cells (2), largely mediated by the downstream mediators RANK-L and WNT4 (3, 6, 7).

High mammographic density (HMD) is another important risk factor of breast cancer (8, 9). Therefore, the architecture and the crosstalk between the stroma including the extracellular matrix (ECM) and epithelial cells in the context of progesterone exposure, play a vital role in breast cancer initiation process.

The mitogenic action of progesterone can be counteracted by PR modulators. There is promising but limited data suggesting the potential efficacy of antiprogesterone in breast cancer prevention (10, 11). Nevertheless, numerous studies have investigated the use of a low continuous dose of antiprogesterone in benign gynecological conditions and breast cancer treatment (12) . We hypothesized that antagonizing progesterone signaling may protect against breast carcinogenesis, warranting further clinical and molecular investigations. As a secondary outcome of our randomized controlled trial (RCT) (13), we studied the effects of the PR-antagonist mifepristone on normal breast tissue following two months of treatment in healthy premenopausal women.

## Materials and methods

This study reports the secondary outcome of a prospective, double blind, placebo-controlled RCT with the main outcome to study the impact of pretreatment with a continuous low dose of mifepristone on menstrual bleeding patterns in women opting for a levonorgestrel releasing intrauterine device for contraception where the results are published by our group elsewhere (13). The secondary outcome presented here, was to investigate the effect of mifepristone treatment on breast tissue. The study was conducted at Karolinska University Hospital, Stockholm, Sweden from 2009 until 2015 and was approved by the Swedish Medical Products Agency (EudraCT number 2009-009014-40). The study protocol was designed according to the recommendations in the CONSORT statement and was approved by the ethical committee at Karolinska Institutet (Dnr: 2009/144-31/4) prior to recruitment. The trial was registered at clinicaltrials.gov (NCT01931657).

### Subjects

Eligible study subjects were healthy premenopausal women aged 18-43 years with regular menstrual cycles lasting 25-35 days and with no contraindications to any of the study treatments. All exclusion criteria are presented in the original study, including use of any hormonal or intrauterine contraception and pregnancy, or breastfeeding two months prior to the study or a history of breast cancer or other malignancies. The trial chart explains the details of the enrolled subjects in the current cohort (Figure S1) and baseline characteristics of the women contributing to paired breast biopsies analyses are presented in Table S1.

### Treatment

Study subjects were randomized into two treatment groups (13). One group was treated with 50 mg mifepristone (one quarter of 200 mg Mifegyne®, Exelgyn) every other day for two months (56 days) starting on the first day of the menstrual cycle. The comparator group received visually indistinguishable B-vitamin tablets (TrioBe® Recip) which were also divided into four parts. For the purpose of the present study, only paired breast samples from the mifepristone treated group were analyzed.

### Biopsy collection

Core needle breast aspiration biopsies were collected at baseline and at the end of the treatment, under ultrasound guidance from the upper outer quadrant of one breast using a 14-gauge needle with an outer diameter of 2.2 mm. The collected breast tissue was divided into two parts, snap-frozen and stored at −180°C until further processing.

In order to do a functional validation with an *in vitro* experiment, we used breast tissue samples from three additional healthy and premenopausal women undergoing mammoplasty procedure. This collection was performed according to a separate study, approved by the ethical committee at Karolinska Institutet (Dnr: 2021-04144) prior to recruitment.

### RNA extraction

RNA extraction was performed from 16 paired samples (i.e. 32 samples) using the Purelink^TM^ RNA Micro kit in conjunction with TRIzol reagent (Life Technologies). For in vitro studies, the RNA extraction from primary breast cells was performed using Quick-DNA/RNA^TM^ Microprep Plus; Zymoresearch. RNA quantification was done using the Qubit RNA High Sensitivity Assay Kit (Invitrogen/ Thermo Fischer).

### cDNA library construction and sequencing

Complementary DNA (cDNA) libraries for NGS were constructed from RNA for the 16 paired samples, before and after mifepristone treatment, using the well-established Smart-seq2 protocol (14), and been well optimized in our group (15).

### RNA-sequencing data processing and analysis

Quality check of raw sequencing reads was done with FastQC and MultiQC (16). RNA sequencing (RNA-seq) data analysis was performed with the Partek Flow Genomic Analysis Software (Partek Inc., St. Louis, Missouri, USA). The detailed analysis optimized by our group is mentioned in von Grothusen et al (15)

### Gene ontology and pathway analysis

GO analyses for the functional annotation of the DEGs and enriched pathway analysis were conducted using the g:Profiler database (version e101_eg48_p14_baf17f0) with Benjamini-Hochberg FDR multiple testing correction method applying significance threshold of 0.05 (18). Reactome pathway analysis (19) and Metascape-designet database (20) were also used.

### RT-PCR analysis

The extracted RNA samples were converted to cDNA using SuperScript^®^ VILO^TM^ kit (Invitrogen®, Thermo Fisher Scientific, Waltham, USA). We validated technically some significantly altered genes obtained by sequencing using Taqman® gene probes namely *CCL18* (assay ID: Hs00268113_m1), *MMP2* assay ID: Hs01548727_m1), *COL1A1* (assay ID: Hs00164004_m1), *COL1A2* (assay ID: Hs01028956_m1), *COL3A1* (assay ID: Hs00943809_m1), and *18s* (4319413E) as housekeeping gene (Thermo Fisher Scientific, Walthem, USA). We designed a customized primer sequence for ADAMTS2-1 (Forward primer sequence ‘cctgacaacccctacttttgc’; reverse primer sequence ‘tgaggatgtcaggtgtcagc’) and performed RT-PCR using Sybr-green PCR assay. 20ng of cDNA from 10 samples was used in triplicates in the RT-PCR and analyzed on a One Step Plus Real-time PCR system (Applied Biosystems, USA) according to the manufactures protocol. Fold change was calculated using the comparative Ct-method. A paired t-test compared the pre- and post-mifepristone treatment groups. To assess the impact of different doses of mifepristone on breast cells in vitro, a two-way ANOVA test was applied after the square root transformation. Significance was considered at a P-value < 0.05. GraphPad Prism 9.1.2 (GraphPad Software Inc., USA) was utilized for the statistical analyses.

### In vitro validation via primary epithelia cell isolation

After breast tissue collection and examination by pathologist, the sample was transferred for tissue digestion and single cells isolation. In brief, the tissue was diced on ice using BSS and DMEM with HEPES. The small tissue fragments were transferred to a mixture of digestion enzymes (hyaluronidase and collagenase 1) and incubated on a rotator at 37°C for 4-18 hours. The digested tissue was then filtered through a 100 μM strainer, and the resulting flowthrough was cultured using a cocktail of Epicult and mammary media.

### Immunofluorescence

Following cell isolation, a total of 50,000 cells were seeded per well in 8-well NuncLab-Tek Chamber Slides (Sigma) and incubated for 72 hours at 37°C in a 5% CO2 incubator. Subsequently, the cells were fixed with 4% paraformaldehyde for 15 minutes at room temperature and then blocked with 2% BSA and 0.1% Triton-X (Sigma) in PBS for 30 minutes at room temperature. Subsequently, the cells were incubated with primary antibodies (CD49f, Rat, #MA5-16884, ThermoFisher Scientific; EPCAM, Goat, R and D Systems Cat# AF960, RRID:AB_355745; CK8, Mouse, Santacruz, sc-8020; CK14, Rabbit, Invitrogen, # MA5-32214) in 2% BSA in PBS for 1 hour at RT. After three washes with PBS, the cells were incubated with the respective secondary antibodies (Alexa Fluor; Donkey anti-Rat 488, Donkey anti-Goat 594, Donkey anti-Mouse 488, and Goat anti-Rabbit 594) in 2% BSA in PBS for 30 minutes at RT. Finally, the samples were mounted using ProLong Gold Antifade Mounting Medium with DAPI.

### In vitro drug treatment assay

Primary isolated breast epithelial cells were cultured at a density of 5 x 10^4^ cells per well in a 12-well plate. On the following day, these cells were subjected to different concentrations of mifepristone treatment (0, 5, 50, and 100 μM) for a duration of three days. Subsequently, the cells were collected, and their lysates were prepared at three distinct time points: the baseline before the initiation of treatment, after one-day of treatment, and upon the completion of the three-day treatment period. To ensure robustness and reliability, we employed cells from three different donors for this study. For each experiment, we conducted three independent replicates.

### In silico data analysis

We employed the R2 Genomics Analysis and Visualization Platform (21) for conducting comparative transcriptomic analysis. This online resource facilitated the examination and assessment of the enrichment gene signature within the breast cancer cohort obtained from TCGA. The user-friendly interface and comprehensive tools provided by R2 allowed for efficiently exploring and statistically validating each gene within the enriched gene cohort. Our approach involved evaluating the differences between normal breast tissue and breast cancer, along with an exploration of their correlation with PR expression and signaling. To ensure statistical robustness, we applied a T-test with false discovery rate correction as a multiple testing approach, employing a P-value cutoff of 0.05.

## Results

### Modulation of gene expression by mifepristone enriches ECM signaling pathways in normal breast tissue

We compared the gene expression profile in normal breast tissue before and after mifepristone treatment. A false discovery rate (FDR) of ≤0.05 and fold change (FC) of ≥2 or ≤-2 was considered statistically significant and identified 27 differentially expressed genes (DEGs) of which 19 genes were upregulated and 8 genes were downregulated (Table S2). We grouped the 27 DEGs and named them Gene signature Enriched to Mifepristone’s action on normal Breast (GEM-B). GEM-B represents a set of genes responsive to mifepristone in normal breast tissue. A volcano plot displaying GEM-B among overall gene expression is presented in Figure S2A. To technically validate RNA-seq data at the individual gene level, we employed real-time (RT)-PCR on the same RNA extracted samples for several genes from the GEM-B (Figure S2B). In line with the transcriptome analysis, the mifepristone treated samples exhibited a significant upregulation of the six validated genes by RT-PCR.

To explore the biological context of the DEGs, gene functional enrichment analysis were performed using the g:Profiler database. The results of the top five terms in each of the three Gene Ontology (GO) categories (BP: biological process; CC: cellular component, MF: molecular function) annotated in the database are presented in Table 1. The Reactome pathway analysis demonstrated the upregulated DEGs significantly enriched in 54 biological processes. The top ten of those, were mainly associated with ECM organization (Table S3). The same analyses with the downregulated DEGs revealed one significantly enriched term in the GO functional annotation (ontology: MF), namely ‘active borate transmembrane transporter activity’ with the involvement of solely gene SLC4A1. In the Reactome pathway analysis, there were two genes involved separately in six pathways; gene IL1B (involved in ‘CLEC7A/inflammasome pathway’, ‘Interleukin-1 processing’, ‘cell recruitment’ and ‘purinergic signaling in leishmaniasis infection’ and gene LAMA1 (involved in ‘laminin interactions’ and ‘MET activates PTK2 signaling’.

**Table 1.**
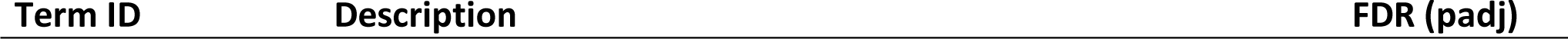
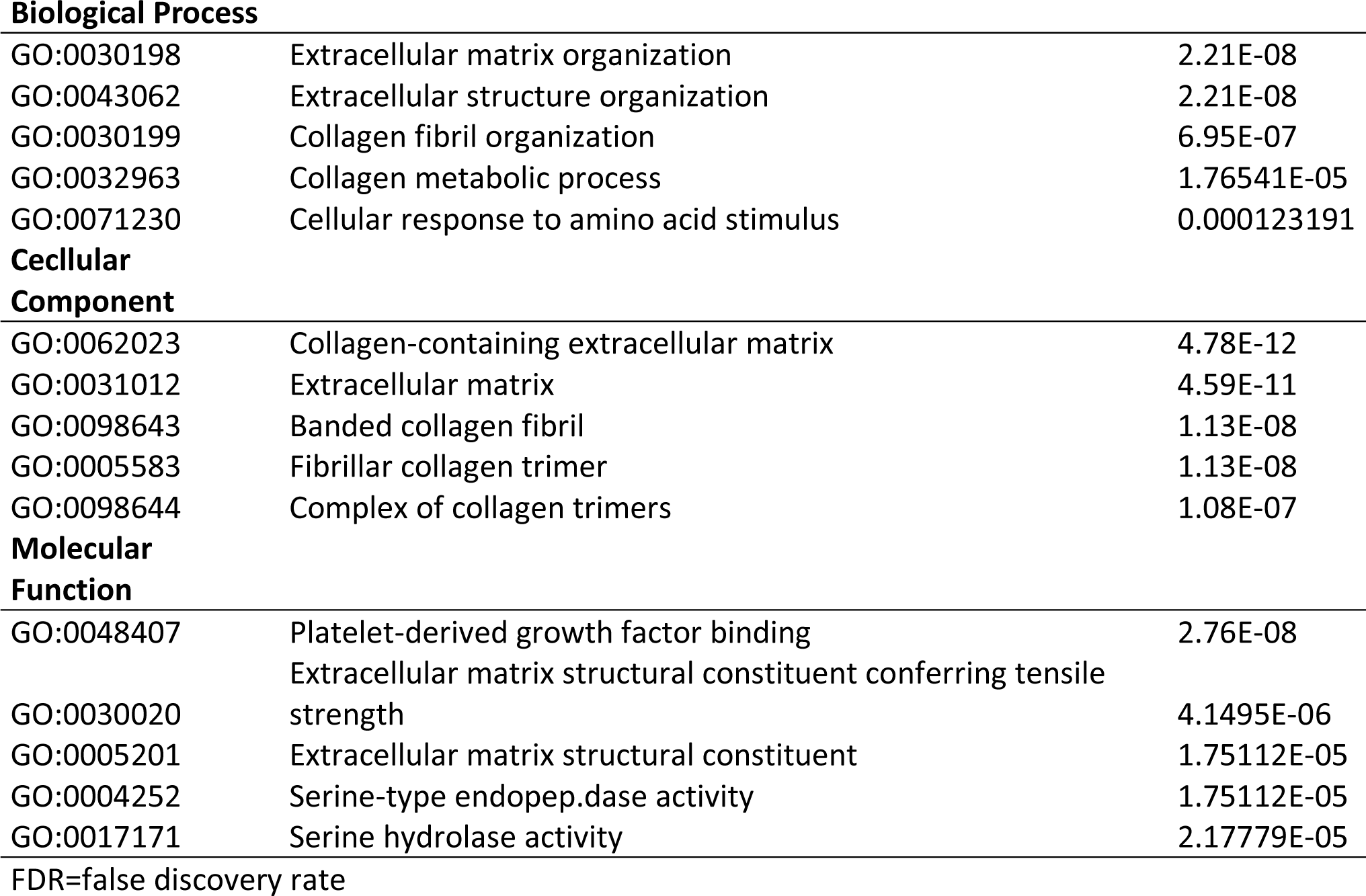
Top 15 enriched gene ontology terms of differentially expressed upregulated genes as determined by Gene Ontology (GO) analyses.

| Term ID | Description | FDR (padj) |
| --- | --- | --- |
| **Biological Process** |  |  |
| GO:0030198 | Extracellular matrix organization | 2.21E-08 |
| GO:0043062 | Extracellular structure organization | 2.21E-08 |
| GO:0030199 | Collagen fibril organization | 6.95E-07 |
| GO:0032963 | Collagen metabolic process | 1.76541E-05 |
| GO:0071230 | Cellular response to amino acid stimulus | 0.000123191 |
| **Cellular****Component** |  |  |
| GO:0062023 | Collagen-containing extracellular matrix | 4.78E-12 |
| GO:0031012 | Extracellular matrix | 4.59E-11 |
| GO:0098643 | Banded collagen fibril | 1.13E-08 |
| GO:0005583 | Fibrillar collagen trimer | 1.13E-08 |
| GO:0098644 | Complex of collagen trimers | 1.08E-07 |
| **Molecular****Function** |  |  |
| GO:0048407 | Platelet-derived growth factor binding | 2.76E-08 |
| GO:0030020 | Extracellular matrix structural constituent conferring tensile strength | 4.1495E-06 |
| GO:0005201 | Extracellular matrix structural constituent | 1.75112E-05 |
| GO:0004252 | Serine-type endopeptidase activity | 1.75112E-05 |
| GO:0017171 | Serine hydrolase activity | 2.17779E-05 |

FDR=false discovery rate

The analysis was validated via a third database, Metascape-designet database. The data set enrichment from designet pathway indicated similarly the enrichment of ECM-related pathways (Figure S3).

### The in vivo effect of mifepristone is comparable to its in vitro effect on normal breast tissue

To further validate the *in vivo* changes in the transcriptomic signature induced by mifepristone treatment, we isolated primary breast epithelial cells and exposed them to varying concentrations of mifepristone (0, 5, 50, and 100 μM) during two different treatment periods (one and three days), to study the dose response effect as well as the influence of duration of treatment.

First, we characterized the primary isolated cells and assessed the enrichment of distinct epithelial cell subtypes including luminal progenitor, mature luminal, basal, and other stromal cells. The expression of four protein markers (EPCAM, CD49f, CK8, and CK14) was examined (22). Notably, we identified diverse expression patterns; some cells exhibited positivity for CD49f or CK14, indicating a basal phenotype. Mature luminal cells expressed EPCAM or CK8. In contrast, other cells were positive for both CD49f and EPCAM, suggesting a luminal progenitor phenotype (Figure 1A). These findings underscore the significance of the heterogeneity within the isolated cells, emphasizing the need to capture the holistic impact of the drug during *in vitro* treatment.

**Figure 1.**
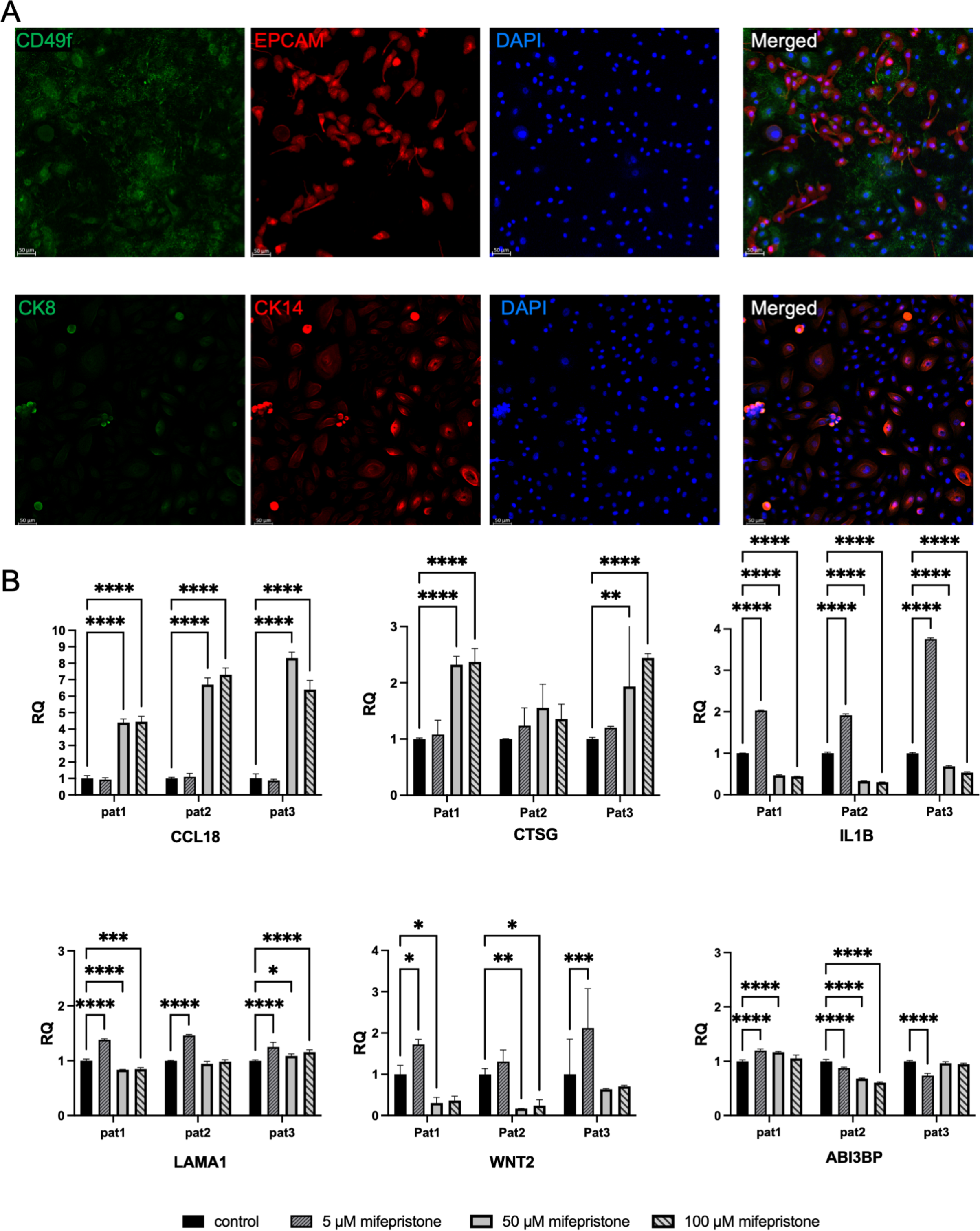
**A)** Characterization of normal primary breast cell (Basal, luminal progenitor & Mature cells). The upper panel displays immunofluorescent images depicting breast cells stained with CD49f (green) and EPCAM (red), while the lower panel showcases cells stained with CK14 (red) and CK8 (green). Scale bars indicate 50 µm. DAPI was used to detect the nuclei. **B)** Assessment of relative gene expression in normal primary breast cells. Real time PCR analysis was conducted on selected genes (CCL18, CTSG, ABI3BP, WNT2, IL1b, and LAMA1) to evaluate their expression levels in normal primary breast cells treated with varying concentrations of mifepristone (control, 5μM, 50μM, 100μM) for a three-day duration. Statistical significance is denoted as follows: *p<0.0001; **p<0.01; ***p<0.001; ****p<0.0001.

Based on our analysis of RNA-seq data from the *in vivo* clinical trial, six candidate genes were selected among the top upregulated and downregulated ones (CCL18, CTSG, ABI3BP, LAMA1, IL1b and WNT2) (Figure 1B and Figure S4). Consistent with our findings from the RNA-seq data, the expression levels of CCL18 and CTSG were upregulated in the *in vitro* experiment after longer treatment with higher concentrations, and the same was observed following shorter treatment; except for one patient where CTSG expression showed a non-significantly upregulated trend and remained unaffected, respectively. WNT2 demonstrated upregulation in all patients when treated with low doses over a three-day period. Conversely, it exhibited downregulation at higher concentrations during both treatment periods. Notably, ABI3BP demonstrated considerable inter-patient variability; it was both up- and downregulated following different concentrations seen in both treatment periods. LAMA1 was not aligned with its downregulated pattern seen in the RCT cohort; it was mainly upregulated following treatment *in vitro*. However, the secondly most downregulated DEG, IL1B, was significantly reduced with higher concentrations of mifepristone following longer treatment and the same was seen for two patients following shorter treatment. Interestingly, IL1B showed a significant upregulation with low doses of mifepristone as compared to the untreated cells.

### GEM-B is associated with breast carcinogenesis

Given the recognized protooncogenic effect of progesterone in breast carcinogenesis, we aimed to explore the enrichment of the GEM-B signature within breast cancer samples. To achieve this objective, we systematically examined the expression patterns of our GEM-B signature in the breast cancer dataset from The Cancer Genome Atlas (TCGA) using the R2 platform.

The signature was examined comparing the RNA-seq data of primary breast cancer tissue (n=1101) to normal breast tissue (n=113). The results revealed a notable enrichment of GEM-B in the TCGA data cohort when comparing cancerous to normal tissue. Specifically, 21 out of the 27 signature genes exhibited significant and differential expression when comparing breast cancer to normal breast tissue.

However, this enrichment displayed a dichotomy between cancerous and normal tissue. Out of the 21 significantly correlated genes, 11 were enriched in normal breast tissue compared to tumor (ABI3BP, CTSG, DPP4, CCL18, OSR2, GRIA3, MMP2, LAMA1, ASPRV1, IL1B, PRR4), while the remaining 10 genes were significantly enriched in tumor tissue compared to normal (COL1A1, COL5A1, COL1A2, COL3A1, WNT2, C1QTNF3, ADAMTS2, GXYLT2, CCDC157, RP1), shown in Figure 2. Our findings underscore a substantial correlation between GEM-B, comprising approximately 77.7% of the signature (21 out of 27 genes), and breast carcinogenesis.

**Figure 2.**
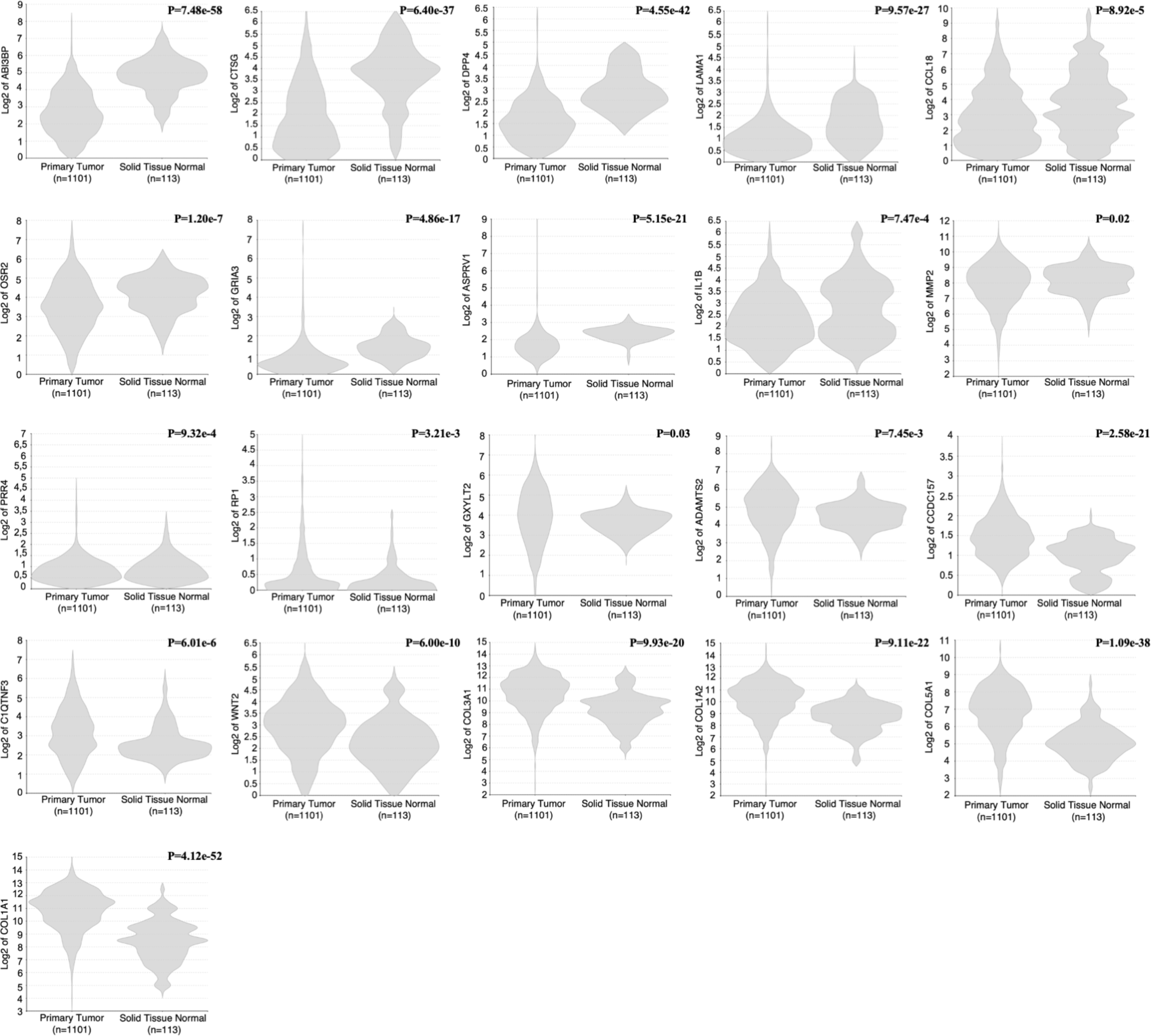
GEM-NB signature enrichment in breast cancer. Utilizing the R2 platform and the TCGA breast cancer dataset, the expression patterns of each gene within the GEM-NB signature were examined. Comparative analysis involved RNA-seq data between two the groups of primary breast cancer tissue (n=1101) and normal breast tissue (n=113).

### GEM-B is significantly correlated to PR expression status in breast cancer

Having investigated the impact of mifepristone on healthy women, we aimed to explore the potential relevance of the transcriptomic changes resulting from mifepristone treatment in our cohort on PR status of the breast cancer cohort. The TCGA breast cancer dataset facilitated the stratification of the PR status across the entire cohort, with 777 patients classified as PR-positive and 337 patients as PR-negative. We conducted an in-depth analysis of the GEM-B gene list, with a focus on delineating the PR status distinctions within the breast cancer datasets.

Out of the 27 genes within the GEM-B signature, 20 exhibited significant enrichment when comparing the PR status categories. Notably, the majority of the enriched genes demonstrated a robust correlation with PR expression. Specifically, 17 out of the 20 enriched genes (TPSAB1, TPSB2, C1QTNF3, PIEZO2, CTSG, COL1A2, COL1A1, COL3A1, OSR2, ZNF620, ABI3BP, MMP2, GXYLT2, COL5A1, CCDC157, RP1, ADAMTS2) exhibited higher expression levels in PR-positive cancer tissue compared to PR-negative counterparts, while only 3 genes (CCL18, SLC4A11, LAMA1) displayed heightened enrichment in PR-negative tissue (Figure 3).

**Figure 3.**
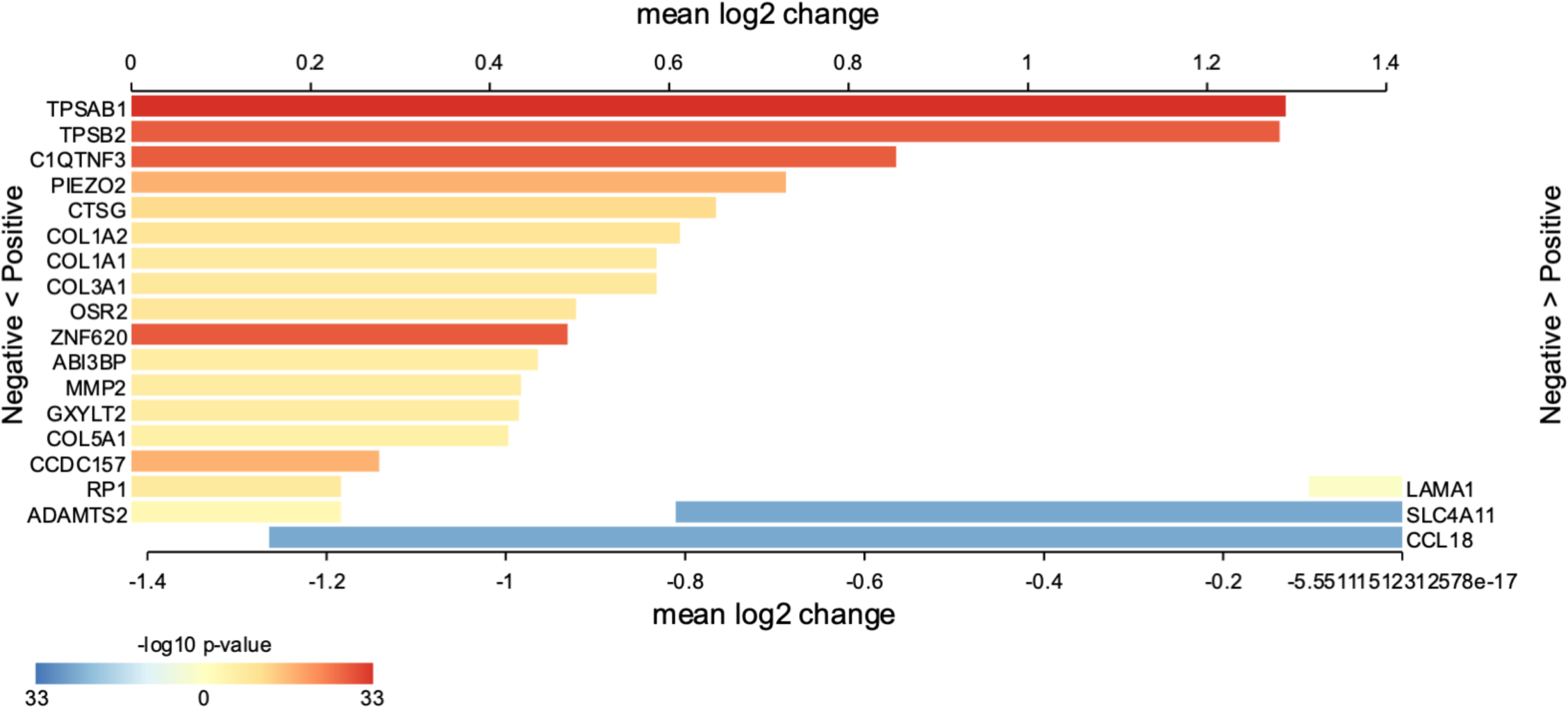
Mifepristone driven transcriptomic changes in the context of PR status in breast cancer. Employing the R2 platform and the TCGA breast cancer dataset, seventeen genes within the GEM-NB signature show significant enrichment in PR positive cohort, whereas three genes were enriched in PR negative cohort. The results are presented as a heatmap illustrating the mean log2 values for each gene, with P values indicated on a log10 scale. The analysis involved RNA-seq data from 1114 breast cancer tissues, comparing PR-positive cases (n=777) to PR-negative cases (n=337).

## Discussion

In the present study, transcriptomic profiling and subsequent bioinformatics analyses were performed to explore gene expression response associated with antagonizing progesterone in breast tissue of healthy premenopausal women. The study focuses on role of endogenous progesterone, enlightening the impact of mifepristone in driving the gene expression patterns and ECM signaling pathways. The findings indicate a correlation with breast carcinogenesis and PR expression status in breast cancer.

The cycle driven intermittent progesterone exposure followed by mammary gland regression have been emphasized as important causes of tumorigenesis, as opposed to gradual and continuous elevations during pregnancy or anovulation including lactational amenorrhea (1, 2). RNA-seq from normal breast tissue in the Komen bank allies with progesterone’s mitogenic role. About 87% of the upregulated genes in the luteal phase, emphasize paracrine action of RANKL, WNT4, and epiregulin as well as enriched functions of DNA replication, mitosis, and DNA repair (23).

To address rising breast cancer incidence (8), exploring novel preventive agents is crucial. Mifepristone, a widely studied PR modulator in various benign gynecological conditions and breast cancer inhibition (12), may also hold potential in breast cancer prevention. In a rodent model, mifepristone had a reverse effect on murine mammary stem cell expansion and progesterone’s paracrine effectors (24), although caution is needed in translating animal-based results to human *in vivo* conditions.

Only two placebo-controlled trials have assessed the effect of mifepristone in normal premenopausal human breast tissue *in vivo*. Exposure to mifepristone for two or three months, significantly reduced Ki-67 expression in breast tissue (10), suggesting inhibition of breast epithelial cell proliferation, and decreased mitotic age surrogate marker and luminal progenitor cell fraction in all analyzed healthy controls (11).

Our findings highlight the enrichment of several pathways that directly regulate and drive extracellular structure organization and function. ECM displays a pivotal role in tissue homeostasis; consequently, dysregulation and destruction of ECM dynamics can lead to tumorigeneses and cancer development (25, 26). During the menstrual cycle, it undergoes hormonal regulation, affecting cell signaling and cancer pathways in the mammary gland and the surrounding microenvironment (2). Clinically, HMD is positively associated with collagen, ECM density, and the epithelial and stromal compartments, but negatively with fat tissue (8, 9). One of the main structural ECM proteins are collagen which represent a key factor that provide tensile strength to the ECM (26) and in the present study, different collagens (COL1A1, COL1A2, COL3A1, COL5A1) were significantly enriched upon mifepristone treatment. Moreover, both collagen degradation and formation emerged as enriched pathways in our material, reflecting an increased remodeling of the ECM compared to baseline. These findings may reflect an ongoing adaptation to mifepristone and a longer treatment protocol might have revealed the eventual direction in which the equilibrium would shift. Nevertheless, it seems that the regulation of ECM plays a central role in progesterone action and progesterone receptor antagonism in the breast.

A fundamental approach for assessing progesterone signaling involves assessment of the PR expression and the subsequent downstream actions within the signaling pathway. The notable enrichment of 20 genes within the GEM-B signature in PR-positive breast cancer tissue underscores the substantial involvement of progesterone in the development of breast cancer. Furthermore, our findings suggest that blocking progesterone signaling by mifepristone may play a significant role in preventing the initiation of breast cancer. This prompts further avenues for in-depth mechanistic studies, shedding light on the potential of mifepristone as both a preventive and therapeutic strategy for breast cancer.

To the best of our knowledge, the present study is the first to explore the changes in the transcriptomic landscape and its biological functions following progesterone antagonism with mifepristone treatment in healthy breast tissue *in vivo*. The sample population originates from a double-blind RCT (13), limiting the bias in the results and individual paired samples were used, thus reducing the inter-individual variability. Next Generation Sequencing (NGS) was used to identify DEGs, and the results were validated with RT-PCR, confirming the expression pattern for all six randomly chosen genes; reinforcing that data derived from RNA-seq technology is of robust nature and could be applied for further analyses. However, long-term effects after treatment discontinuation were not elucidated due to lack of follow-up data. Based on indications of ECM remodeling following mifepristone treatment, measurements of breast stiffness and density through mammographies could provide further insights. Even though breast cancer seems to arise predominantly from epithelial cells (2), the stroma of mammary gland comprised mainly of the ECM emerged in our study to play a key role which is in line with a plethora of investigations on breast cancer, even suggesting ECM remodeling as a potential therapeutic target (26).

In conclusion, our investigation into PR modulation in normal breast tissue, following mifepristone treatment, uncovers crucial alterations in gene expression patterns. The observed shifts in gene expression, in pathways related to ECM organization, point to the complicated involvement of ECM dynamics. The significant correlation of our enriched signature with the PR expression in breast cancer emphasizes the downstream impact of progesterone. Undoubtedly, comprehensive studies specifically designed to delve into the detailed molecular landscape alterations induced by mifepristone treatment could shed light into the molecular actions of progesterone. Furthermore, such studies may explore whether antagonizing progesterone could accord protective properties in the breast.

## Data Availability

The raw data and processed data files were submitted to Gene Expression Omnibus database.

## Acknowledgments

We would like to thank research midwife Eva Broberg for patient recruitment support and Dr Birgitte Wilczek for offering her kind help in collecting breast biopsies. Our thanks also go to Dr. Angelique Flöter Rådestad and Dr. Inkeri Leonardsson Schultz for their contribution in providing breast samples from healthy women who underwent reduction mammoplasty surgeries.

---

**Supplemental figure 1.** Study flow diagram.

*Paired biopsies = baseline plus follow-up breast biopsies from the same patient.

**Supplemental figure 2. A)** Volcano plot showing the distribution of the differentially expressed genes (DEGs) between breast samples from baseline and after mifepristone treatment. X-axis represent log2 fold change and Y-axis represent logFDR (adjusted p-value). Black vertical lines show log fold change of - 2 and 2 while the horizontal black line represents a p-value of 0.05. The points represent genes; red for the upregulated DEGs, blue for the downregulated DEGs and grey for the non-differentially expressed genes. **B)** Assessment of relative gene expression before and after mifepristone treatment. Utilizing the RNA isolated from the breast tissue obtained from RCT, Real time PCR analysis was conducted on selected genes (ADAMTS2-1, CCL18, MMP2, COL1A1, COL1A2 and COL3A1) show a significant upregulation with mifepristone treatment in breast. This is in line with RNA sequencing data. *p<0.05; **p<0.01; ***p<0.001.

**Supplemental figure 3**. Network of enriched terms for GEM-NB genes as determined by Metascape-designet analysis; showing the significance enrichment of extracellular matrix remodeling pathways. (A) Nodes colored by cluster ID; nodes that share the same cluster ID are typically close to each other. (B) Nodes colored by p-value, where terms containing more genes tend to exhibit more significant p-values.

**Supplemental figure 4**. Assessment of relative gene expression in normal primary breast cells. Real time PCR analysis was conducted on selected genes (CCL18, CTSG, ABI3BP, WNT2, IL1b, and LAMA1) to evaluate their expression levels in normal primary breast cells treated with varying concentrations of mifepristone (control, 5μM, 50μM, 100μM) for one day duration. Statistical significance is denoted as follows: *p<0.0001; **p<0.01; ***p<0.001; ****p<0.0001.

**Supplemental table 1**. Baseline characteristics of women contributing to paired breast biopsies, before and after mifepristone treatment, expressed as median (range).

**Supplemental table 2**. Differentially expressed genes with mifepristone treatment.

**Supplemental table 3**. Top 10 enriched pathways of the upregulated differentially expressed genes as determined by Reactome pathway analysis.

