## Supplementary tables and figures for "Transcriptomic profile of normal breast tissue post-mifepristone treatment: secondary outcomes of a randomized controlled trial"

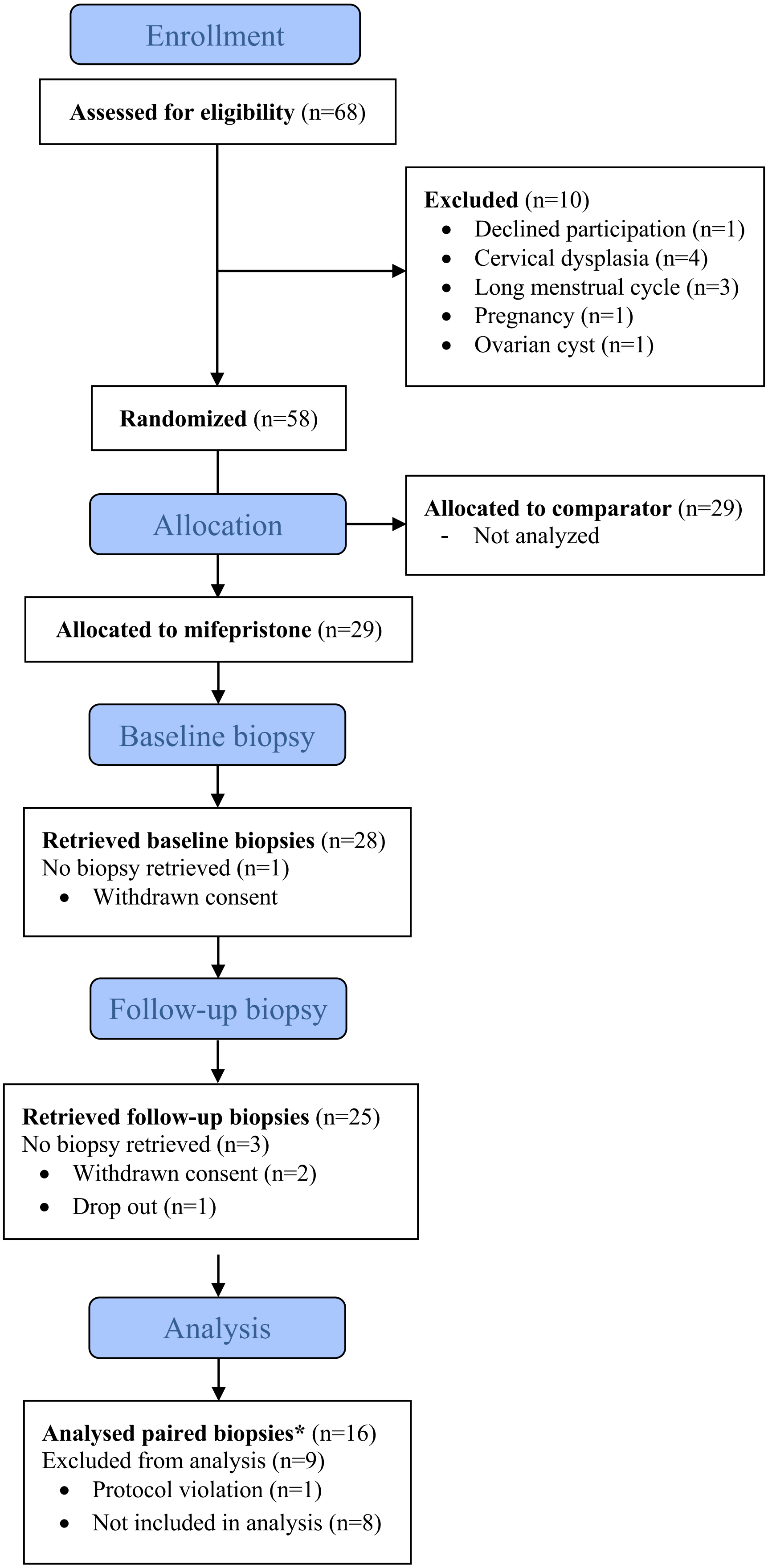

Enrollment
Assessed for eligibility (n=68)
Excluded (n=10)
Declined participation (n=1)
Cervical dysplasia (n=4)
Long menstrual cycle (n=3)
Pregnancy (n=1)
Ovarian cyst (n=1)
Randomized (n=58)
Allocation
Allocated to comparator (n=29)
- Not analyzed
Allocated to mifepristone (n=29)
Baseline biopsy
Retrieved baseline biopsies (n=28)
No biopsy retrieved (n=1)
Withdrawn consent
Follow-up biopsy
Retrieved follow-up biopsies (n=25)
No biopsy retrieved (n=3)
Withdrawn consent (n=2)
Drop out (n=1)
Analysis
Analysed paired biopsies* (n=16)
Excluded from analysis (n=9)
Protocol violation (n=1)
Not included in analysis (n=8)


suppl Fig 1

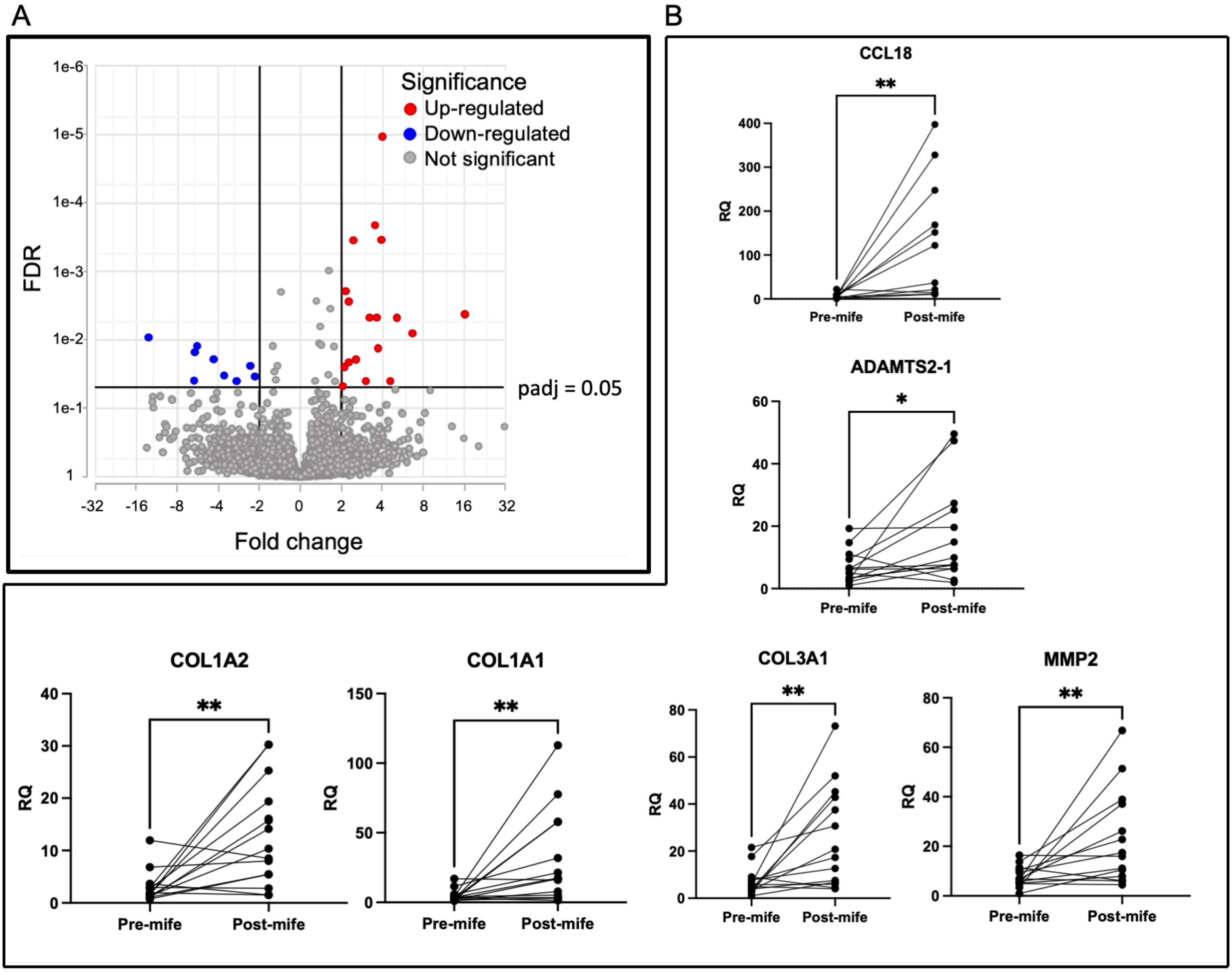

A
B
FDR
Fold change
Significance
Up-regulated
Down-regulated
Not significant
padj = 0.05
1e-6
1e-5
1e-4
1e-3
1e-2
1e-1
1
-32
-16
-8
-4
-2
0
2
4
8
16
32
CCL18
ADAMTS2-1
COL1A2
COL1A1
COL3A1
MMP2
RQ
Pre-mife
Post-mife
**
*


Suppl Fig 2

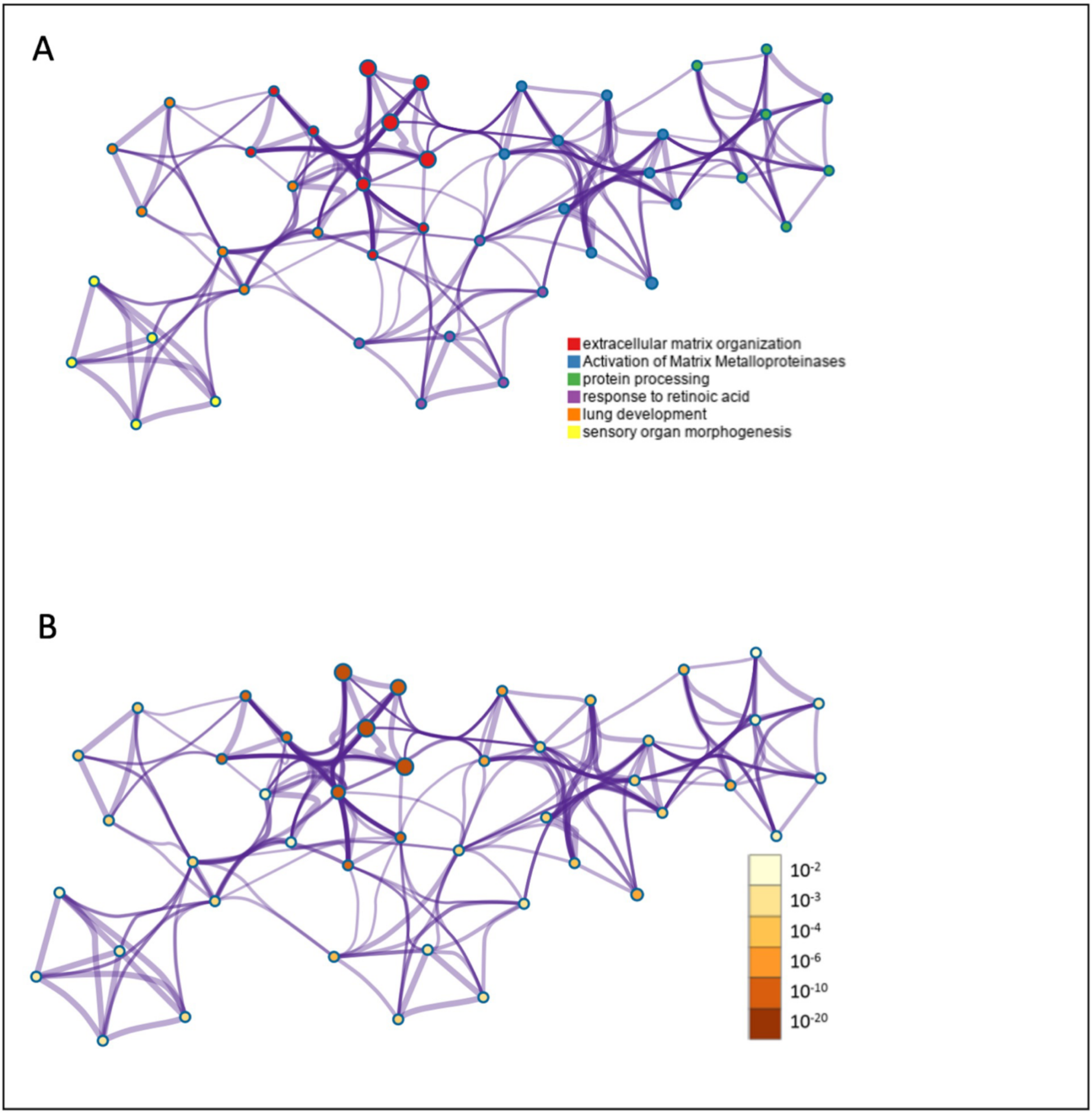
A
extracellular matrix organization
Activation of Matrix Metalloproteinases
protein processing
response to retinoic acid
lung development
sensory organ morphogenesis
B
10-2
10-3
10-4
10-6
10-10
10-20

Suppl Fig 3

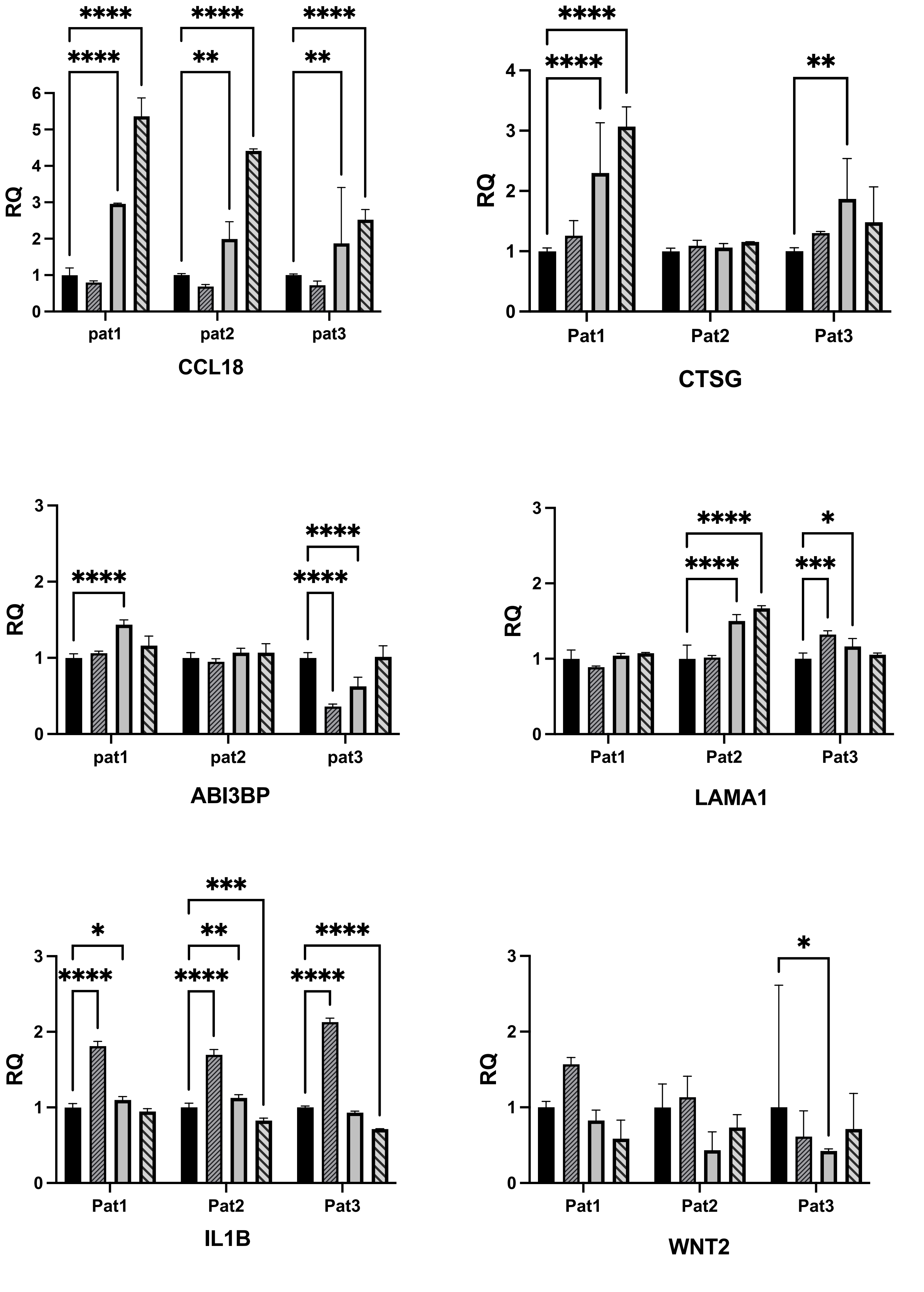
****
****
**
**
6
5
4
3
2
1
0
RQ
pat1
pat2
pat3
CCL18
4
3
2
1
0
RQ
Pat1
Pat2
Pat3
CTSG
*
***
3
2
1
0
RQ
pat1
pat2
pat3
ABI3BP
RQ
Pat1
Pat2
Pat3
LAMA1
RQ
Pat1
Pat2
Pat3
IL1B
RQ
Pat1
Pat2
Pat3
WNT2
control
5 μM mifepristone
50 μM mifepristone
100 μM mifepristone

Suppl Fig 3

**Supplemental table 1**. Baseline characteristics of women contributing to paired breast biopsies, before and after mifepristone treatment, expressed as median (range).

| | |
|---|---|
| Age (years) | 33 (21 – 41) |
| Pregnancies | 1.5 (0 – 5) |
| Parity | 2 (0 – 3) |
| Cycle length (days) | 29 (27 – 32) |
| Menstrual period (days) | 5 (3 – 7) |
| Body Mass Index | 25.8 (18.4 – 32.0) |

**Supplemental table 2. Differentially expressed genes with mifepristone treatment**

| Gene name | P-value | FDR | Fold change |
|---|---|---|---|
| **A. Upregulated genes** | | | |
| CCL18 | 2,41E-06 | 0.0041 | 16.0 |
| WNT2 | 3,40E-06 | 0.0047 | 5.1 |
| CTSG | 7,82E-05 | 0.0397 | 4.5 |
| TPSB2 | 5,50E-10 | 0.0001 | 4.0 |
| TPSAB1 | 6,58E-08 | 0.0003 | 4.0 |
| PIEZO2 | 1,54E-05 | 0.0132 | 3.7 |
| COL1A1 | 3,48E-06 | 0.0047 | 3.6 |
| DPP4 | 2,17E-08 | 0.0002 | 3.6 |
| GRIA3 | 3,44E-06 | 0.0047 | 3.2 |
| C1QTNF3 | 8,00E-05 | 0.0397 | 3.0 |
| COL1A2 | 2,69E-05 | 0.0196 | 2.6 |
| COL3A1 | 2,70E-05 | 0.0196 | 2.6 |
| OSR2 | 7,28E-08 | 0.0003 | 2.4 |
| CPZ | 1,30E-06 | 0.0027 | 2.3 |
| ADAMTS2 | 2,96E-05 | 0.0206 | 2.3 |
| COL5A1 | 5,93E-07 | 0.0019 | 2.2 |
| MMP2 | 3,90E-05 | 0.0245 | 2.1 |
| GXYLT2 | 1,10E-04 | 0.0480 | 2.1 |
| ABI3BP | 1,06E-04 | 0.0477 | 2.0 |
| **B. Downregulated genes** | | | |
| ZNF620 | 6,26E-05 | 0.03 | -2.16 |
| LAMA1 | 3,59E-05 | 0.02 | -2.34 |
| PRR4 | 8,47E-05 | 0.04 | -2.95 |
| ASPRV1 | 5,73E-05 | 0.03 | -3.64 |
| SLC4A11 | 1,38E-05 | 0.01 | -5.68 |
| CCDC157 | 1,83E-05 | 0.02 | -5.96 |
| IL1B | 8,56E-05 | 0.04 | -6.06 |
| RP1 | 7,80E-06 | 0.01 | -13.00 |

FDR=false discovery rate

**Supplemental table 3.** Top 10 enriched pathways of the upregulated differentially expressed genes as determined by Reactome pathway analysis.

| Pathway ID | Name | FDR (Padj) | Genes |
|---|---|---|---|
| R-HSA-1474228 | Degradation of the extracellular matrix | 1.80E-09 | CTSG. TPSAB1. COL1A1. COL1A2. COL3A1. COL5A1. MMP2 |
| R-HSA-1474244 | Extracellular matrix organization | 2.57E-09 | CTSG. TPSAB1. COL1A1. COL1A2. COL3A1. ADAMTS2. COL5A1. MMP2 |
| R-HSA-1650814 | Collagen biosynthesis and modifying enzymes | 9.22E-08 | COL1A1. COL1A2. COL3A1. ADAMTS2. COL5A1 |
| R-HSA-1442490 | Collagen degradation | 9.22E-08 | COL1A1. COL1A2. COL3A1. COL5A1. MMP2 |
| R-HSA-3000170 | Syndecan interactions | 1.76E-07 | COL1A1. COL1A2. COL3A1. COL5A1 |
| R-HSA-1474290 | Collagen formation | 2.30E-07 | COL1A1. COL1A2. COL3A1. ADAMTS2. COL5A1 |
| R-HSA-8874081 | MET activates PTK2 signaling | 2.30E-07 | COL1A1. COL1A2. COL3A1. COL5A1 |
| R-HSA-8875878 | MET promotes cell motility | 6.69E-07 | COL1A1. COL1A2. COL3A1. COL5A1 |
| R-HSA-8948216 | Collagen chain trimerization | 8.81E-07 | COL1A1. COL1A2. COL3A1. COL5A1 |
| R-HSA-3000171 | Non-integrin membrane-ECM interactions | 2.460x10-6 | COL1A1. COL1A2. COL3A1. COL5A1 |

FDR=false discovery rate
